# Development of a knowledge distillation-based rapid prediction model for 7-day adverse prognosis in emergency patients with acute upper gastrointestinal bleeding

**DOI:** 10.64898/2026.11.24.26368922

**Authors:** Zhaofan Li, Lizun Guo, Zefan Jing, Yinyun Yan, Jiayi He, Hongju Xiao, Yihao Zhou, Tingting Xu, Wenli Qiao, Qingyue Duan, Lili Wang, Yugui Zhang, Li Chen

## Abstract

**Background:** To construct a rapid prediction model for 7-day adverse prognosis in emergency patients with acute upper gastrointestinal bleeding (AUGIB) based on feature-constrained knowledge distillation.

**Methods:** A retrospective analysis was conducted on clinical data from 601 AUGIB patients admitted to the Emergency Department of the First Medical Center of Chinese People’s Liberation Army (PLA) General Hospital from December 2023 to January 2025. Modeling was performed in two stages. First, a teacher model was trained using full-dimensional clinical indicators. Second, a student model (rapid prediction model) was developed. Bedside-obtainable features were selected through feature contribution analysis and physician experience, excluding hematological indicators. Knowledge distillation was introduced from the teacher model. The area under the receiver operating characteristic curve (AUC) served as the primary metric to select the optimal model. The SHapley Additive exPlanations (SHAP) algorithm was used for interpretability analysis.

**Results:** Eleven features were selected to build the rapid prediction model. The Light Gradient Boosting Model (LightGBM) model performed best with an AUC of 0.848, accuracy of 0.826, precision of 0.750, recall of 0.732, F1 score of 0.741, and Brier score of 0.128. SHAP analysis showed that Glasgow Coma Scale, age, and sex held the highest predictive value. The model relies only on immediate bedside indicators such as vital signs and comorbidities. This ensures good timeliness. Its AUC was higher than that of pre-endoscopic Rockall score, Shock Index, Age-Shock Index, and Modified Shock Index (P<0.05). Although the difference was not statistically significant, its AUC was higher than that of Glasgow-Blatchford score, AIMS-65, and Modified Early Warning Score (P>0.05). It showed clear advantages in accuracy and precision. Compared to the optimal teacher model, the AUC decreased by only 0.014, maintaining good discriminatory performance.

**Conclusion:** This rapid prediction model possesses the capability for rapid prediction of 7-day adverse prognosis in emergency AUGIB patients while maintaining certain discriminatory performance. It assists emergency physicians in rapid decision-making and resource allocation. This helps improve prognosis and reduce medical costs.

**Author summary:** Acute upper gastrointestinal bleeding (AUGIB) is a time-sensitive medical emergency, where rapid bedside risk stratification is critical for triage decisions, resource allocation, and timely intervention. Existing clinical risk scoring systems, such as the Glasgow-Blatchford Score (GBS) and AIMS-65, rely heavily on laboratory parameters that are not immediately available during the initial emergency assessment, which inevitably delays clinical decision-making. To address this gap, we developed a knowledge distillation-based rapid prediction model using a teacher-student network architecture. A teacher model was first trained on the full spectrum of clinical indicators. Subsequently, a student model (the rapid prediction model) was trained using only 11 bedside-obtainable features — excluding time-consuming hematological parameters — via knowledge distillation using soft labels generated from the teacher model. The LightGBM-based rapid prediction model achieved an AUC of 0.848 and an accuracy of 0.826. It outperformed four conventional clinical scoring systems (PRS, SI, Age-SI, and MSI; DeLong test, P < 0.05). While its AUC was higher than those of GBS, AIMS-65, and MEWS, the model demonstrated prominent advantages in accuracy and precision. Compared with the optimal teacher model, the AUC decreased by only 0.014, demonstrating that the distillation strategy effectively preserved discriminative performance despite substantial feature reduction. SHAP analysis identified the Glasgow Coma Scale, age, and sex as the top three predictive contributors, providing clinically interpretable evidence to support model transparency. The model relies exclusively on immediately available bedside indicators (vital signs and comorbidities), enabling risk assessment to be completed within minutes — a critical advantage in high-volume emergency settings.

## Introduction

Acute Upper Gastrointestinal Bleeding (AUGIB) is characterized by acute onset and rapid progression. It has an annual incidence of 50–150 per 100,000 people, a mortality rate of 2.5–10%, and a recurrence rate of 11.9% [1]. It is one of the common critical illnesses in the emergency department. With advancements in pharmacotherapy and endoscopic diagnosis and treatment, most patients receive timely treatment and have a good prognosis. However, a small number of high-risk patients are still not identified in time, leading to severe consequences. Studies have shown that failure to assess the risk of AUGIB in a timely and accurate manner and act quickly may lead to patient deterioration and even life-threatening conditions [2]. For hemodynamically unstable patients, undergoing endoscopy within 6 – 24 hours is associated with reduced in-hospital mortality [3]. Studies also indicate that for patients with AUGIB and hypovolemic shock, initial resuscitation must be initiated as early as possible [4]. Therefore, implementing rapid and accurate assessment for early adverse prognosis in emergency AUGIB is of great significance. It aids in clinical decision-making, such as initiating effective hemostasis and fluid resuscitation strategies, and reducing mortality.

Currently, multiple expert consensus guidelines recommend using risk assessment systems to predict AUGIB prognosis. This aims to identify high-risk patients, stratify treatment, and improve outcomes [5–8]. However, commonly used clinical risk assessment systems, such as the pre-endoscopic Rockall score (PRS), Glasgow-Blatchford score (GBS), and AIMS-65 score, vary in their predictive efficacy [9,10]. The GBS score performs well in predicting the need for clinical interventions such as blood transfusion, endoscopy, or surgery. It is often used to identify low-risk AUGIB patients [11–13]. The AIMS-65 score is significantly superior to Rockall and GBS in predicting mortality [14–17]. However, a common limitation is their reliance on blood indicators. The time required for laboratory testing leads to low assessment efficiency. The PRS score does not include blood indicators. Although it allows for faster assessment, its poor classification performance has been confirmed by multicenter studies [12]. Emergency departments often face a shortage of medical staff and a large number of patients. There is a mismatch between medical resources and actual needs. This emphasizes the need for rapid triage to shorten the time to intervention. However, there is currently no single risk assessment system that supports rapid and accurate measurement of various adverse prognoses in AUGIB.

Machine learning (ML) technology [18–19] is widely used for prognostic evaluation in AUGIB. Nazarian et al. [20] used five categories of indicators, including age and laboratory results, to establish a random forest model for predicting the need for hemostasis. The model achieved an area under the receiver operating characteristic curve (AUC) of 0.84, outperforming the GBS scoring system (AUC 0.75, cutoff value ≥ 12). Zhuang et al. [21] included indicators such as lactate and albumin to construct a prediction model for rebleeding risk in AUGIB patients, achieving an AUC of 0.983. Regarding mortality risk prediction, Deshmukh et al. [22] confirmed that ML models outperformed the Acute Physiology and Chronic Health Evaluation (APACHE) IV scoring system (AUC 0.85 vs. 0.80). In terms of composite risk prediction, Shung et al. [23] developed a ML model to predict composite endpoints such as transfusion, intervention, or death within 30 days. The AUC reached 0.90, significantly outperforming GBS and AIMS-65 scores. However, the above studies still rely on hematological indicators, resulting in poor timeliness in assessment.

To address the conflict between the urgent need for rapid assessment in emergency AUGIB patients and the time-consuming nature of hematological tests, constructing a rapid prediction model is proposed. This model relies on “immediate information” such as basic demographics, vital signs, major symptoms, comorbidities, and simple scores. It supports composite outcome prediction, which will greatly enhance usability and shorten assessment time. However, directly discarding high-weight hematological indicators to train ML models often leads to a serious loss of valid information. This significantly reduces model performance and robustness. Investigations show that in the field of deep learning, knowledge distillation technology can transfer the generalization ability of complex models (teacher models) to simplified models (student models) [24–27]. It has shown great potential in medical auxiliary diagnosis based on image recognition. The implication for this study is to use a “teacher model” containing multi-dimensional features (including hematological results) to guide the training of a “student model.” The student model only uses clinical immediate information (excluding hematological results). By learning the “soft labels” output by the teacher model, the student model simulates the teacher’s decision process. It also captures potential probability distribution information between data categories, known as “dark knowledge”. Therefore, even with limited input features, the student model can approximate the prediction accuracy of the full-dimensional model to the greatest extent. In view of this, this study proposes to use a knowledge distillation strategy with expert-guided feature constraints. We aim to explore a comprehensive risk assessment system for rapid triage while maintaining certain accuracy. This will provide a decision-making tool for the early management of emergency AUGIB patients.

## Methods

This study proposes a rapid prediction model based on feature-constrained knowledge distillation (Fig 1). First, a high-precision teacher model was constructed using full-dimensional features. Then, soft labels output by the teacher model guided the student model with restricted input features. This allowed the student model to simulate the teacher model’s decision process. Finally, the method’s effectiveness was verified by comparing it with commonly used clinical risk assessment systems.

**Fig 1.**
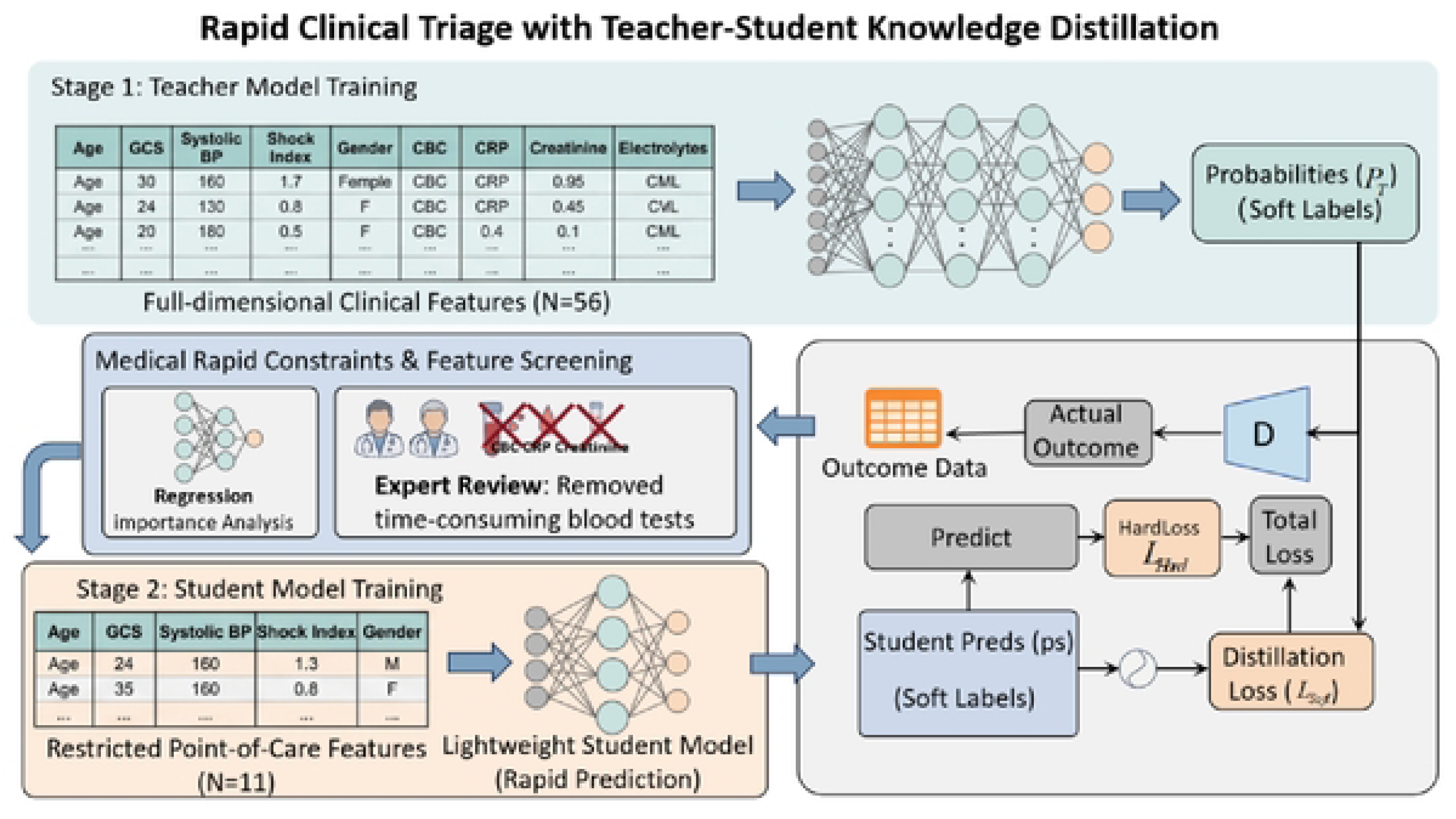
Overall algorithm framework.

### Data source

A retrospective analysis was conducted on 601 AUGIB patients admitted to the Emergency Department of the First Medical Center of Chinese People’s Liberation Army (PLA) General Hospital from December 2023 to January 2025. The data access period is from May 19, 2025 to June 15, 2025. Inclusion criteria were: (1) age ≥ 18 years; and (2) patients meeting one or more of the following conditions according to the “Expert Consensus on Emergency Diagnosis and Treatment Process for Acute Upper Gastrointestinal Bleeding” [28]: definite hematemesis, melena, or hematochezia; positive occult blood in gastric juice or feces; unexplained anemia or unstable vital signs; or endoscopic confirmation. Exclusion criteria were: (1) incomplete key data preventing score calculation; (2) hematological system diseases; and (3) other types of shock.

### Sample size estimation

The 10-fold Events Per Variable (EPV) principle was used to estimate the number of events. We planned to include a maximum of 12 predictor variables. Therefore, the final dataset was required to contain at least 120 events.

### Data collection

1. Clinical indicators (47 items): Based on literature review, basic patient information (excluding any personal identification information, such as name, ID number, phone number, etc.) and physiological/biochemical indicators from the first day in the emergency resuscitation room were collected. Risk assessment system scores were also calculated. The time difference between vital signs collection and blood sampling did not exceed 30 minutes. Blood gas analysis results came from arterial blood, while other results came from venous blood. (2) Primary symptoms (2 items). (3) Comorbidities and underlying diseases (13 items), as detailed in Table 1.The observation period was defined as 7 days after admission to the emergency resuscitation room. Adverse prognosis included death, rebleeding [29], rescue measures (intubation, CPR, defibrillation), emergency intervention, or ICU transfer; otherwise, good prognosis. Early discharge outcomes were confirmed by telephone.

### Data processing

Features with more than 20% missing values were deleted. Missing values for numerical features were filled with the mean. Categorical features underwent label encoding and were filled with the mode (Sex: 0 = female, 1 = male; Primary symptoms, comorbidities, and underlying diseases: 0 = no, 1 = yes). For outlier detection, the Z-score method was used for normally distributed continuous variables (|Z| > 3 was defined as an outlier). For non-normally distributed continuous variables, the interquartile range method was used (values exceeding Q₁ − 1.5 × IQR or Q₃ + 1.5 × IQR were defined as outliers). All outliers were processed using the clipping method.

**Table 1.** Details of collected data.

| Main items | Specific Names |
|---|---|
| **Clinical Indicators**<br>(47) | Gender, Age, Temperature(T), Heart rate(HR), Respiratory rate(RR), Systolic blood pressure(SBP), Diastolic blood pressure(DBP), Power of Hydrogen(PH), Arterial O2 pressure(PO2), Arterial CO2 pressure(PCO2), Base excess(BE), Lactic acid(LA), Alanine aminotransferase(ALT), Aspartate aminotransferase(AST), Albumin(ALB), Total bilirubin(TBIL), Glucose(GLU), Blood urea nitrogen(BUN), Serum creatinine(SCR), Calcium(Ca), Potassium(K), Sodium(Na), Amylase(AMY), Lipase(LIP), Neutrophil count(NEU), White blood cell(WBC), Hemoglobin(HB), Platelet(PLT), Hematocrit(HCT), C-Reactive protein(CRP), Interleukin-6(IL-6), Procalcitonin(PCT), Activated partial thromboplastin time(APTT), Prothrombin time(PT), International normalized ratio(INR), Fibrinogen(FIB), D-Dimer, Lac/Albumin(L/A), Urea/Creatinine(U/C), PRS, GBS, AIMS-65, Glasgow Coma Scale (GCS), Shock Index (SI), Age-Shock Index (Age-SI), Modified Shock Index (MSI), and Modified Early Warning Score (MEWS) |
| **Primary Symptoms**<br>(2) | Hematemesis, Syncope |
| **Comorbidities and Underlying Diseases**<br>(13) | Liver disease(LD), Liver failure(LF), Renal failure, Heart failure(HF), Respiratory failure, Shock, Ischemic heart disease(IHD), Diabetes mellitus(DM), Hypertension(HNT), Coronary heart disease(CHD), Upper gastrointestinal tumor(UGT), Other tumors(OT), and Metastasis of tumor(MOT) |

### Teacher model training

To enhance model interpretability, a ML ensemble was adopted to construct the teacher model. After validating and selecting the optimal model, soft labels containing prior knowledge were generated. These labels guided the student model learning and improved its prediction capability.

First, the collected full-dimensional physiological tabular data *X_Teacher_ = {x_1_, x_2_, …, x_NT_} ∈ R^NT^* was prepared, where *N_T_* represents the quantity of collected physiological data. Based on the concept of knowledge distillation, the full-feature data *T* was input into the teacher model ensemble:

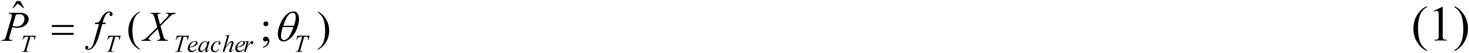

Where *θ_T_* represents the parameters to be optimized in the teacher model, and ^^^ ∈[0,1]^2^ is the probability of predicting adverse prognosis in patients with AUGIB. To better utilize the knowledge extracted by the teacher model, a decoding structure *f_D_* was added. This structure extracted the importance of each indicator to guide the rapid prediction model construction:

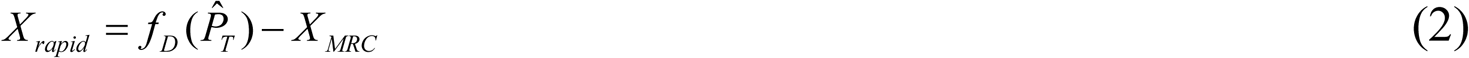

This study used interpretable regression models for feature screening. First, univariate analysis was conducted to screen variables related to the dependent variable. Variables with *P* < 0.1 [30] were retained as candidate independent variables. Second, collinearity diagnostics were performed. Variables with tolerance < 0.1 or variance inflation factor > 5 were excluded. Multivariate Logistic regression analysis was performed on the remaining variables. SPSS used the “Forward: Conditional” stepwise method to build the model. Variables that maximized model fit were introduced stepwise. These variables served as rapid test candidate indicators distilled by the teacher model. Subsequently, Medical Rapid

Constraints *^X^MRC* were added. Two senior emergency physicians removed hematological indicators based on medical experience and knowledge. They also referred to clinical risk assessment systems to supplement non-hematological indicators strongly related to the prediction task. Then, candidate indicators were screened using the aforementioned method. Finally, key features *X_rapid_ = {x_1_, x_2_, …, x_m_} ∈ R^m^* included in the student rapid prediction model were confirmed.

### Student model training based on rapid test feature constraints

First, the rapid test indicators jointly screened by the teacher model and medical prior knowledge in Section 2.1 were used as input features for the student model. Subsequently, the student model was made to learn the soft labels output by the teacher model to capture latent distribution information between data classes.

This study defined the key rapid test features screened in Section 2.2 as set *X_rapid_* = {*x*_1_, *x*_2_,…, *x_m_*}. The student model *S* aimed to establish a non-linear mapping from the low-dimensional feature space to the adverse prognosis risk space:

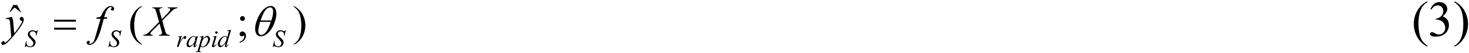

Here, *θ_S_* represented the parameters of the student model to be optimized. To verify the universality and robustness of the model architecture, the student model was also constructed using a ML ensemble, focusing on the introduction of a knowledge distillation strategy in the training process.

In traditional supervised learning, models are guided only by Hard Labels (i.e., confirmed results *y* ∈{0,1}). This causes the model to lose “gray information” of samples in binary classification. To compensate for the fine-grained information loss caused by the absence of hematological indicators in the student model, this study introduced Response-based Knowledge Distillation. After the teacher model *f_T_* was trained on the full-dimensional feature dataset, for any input sample *i*, its output was not just a classification result. It was a probability distribution vector *P_T_* (*x_i_*) containing rich “Dark Knowledge”. This probability value reflected the confidence of the teacher model regarding the adverse prognosis risk. For example, for a sample labeled negative, the teacher model might predict an adverse prognosis probability of 0.45 (high-risk negative) or 0.05 (low-risk negative). Such subtle inter-class correlations cannot be provided by hard labels. Therefore, the total loss function *L_Total_* of the student model was defined as a weighted combination of the true label loss and the distillation loss:

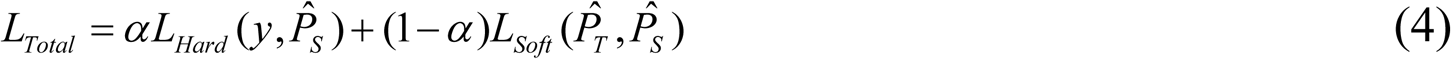

where:

*y* is the clinical Ground Truth;

*P^_s_* is the probability distribution predicted by the student model;

*P^_T_* is the soft probability distribution predicted by the teacher model;

*L_Hard_* utilizes standard Cross-Entropy Loss to ensure the classification accuracy of the student model;

*L_Soft_* utilizes Mean Squared Error (MSE) to force the student’s probability output to approximate the teacher’s, thereby inheriting the teacher’s generalization boundaries in the full feature space;

*α* is a balancing coefficient used to regulate the weight between hard and soft knowledge.

Through this constrained joint training mechanism, the student model *f_S_* not only learns explicit rules (e.g., “Feature A is strongly correlated with adverse prognosis”) but also implicitly internalizes the deep logic of the teacher model *f_T_* (e.g., “Despite the lack of hematological Indicator B, the combination of Vital Sign C and the teacher’s decision process suggests this sample should be classified as high risk”), thereby significantly enhancing the prediction performance.

### Experimental preparation

Based on the data collected in Section 1.2, 56 full-dimensional features were selected to train the teacher model. These features included basic information, vital signs, and hematological results. IL-6 and PCT were excluded because they had more than 20% missing values. Subsequently, feature distillation was performed using the teacher model constructed in Section 2.1. This process yielded 11 candidate rapid test features (details shown in Fig 2).

**Fig 2.**
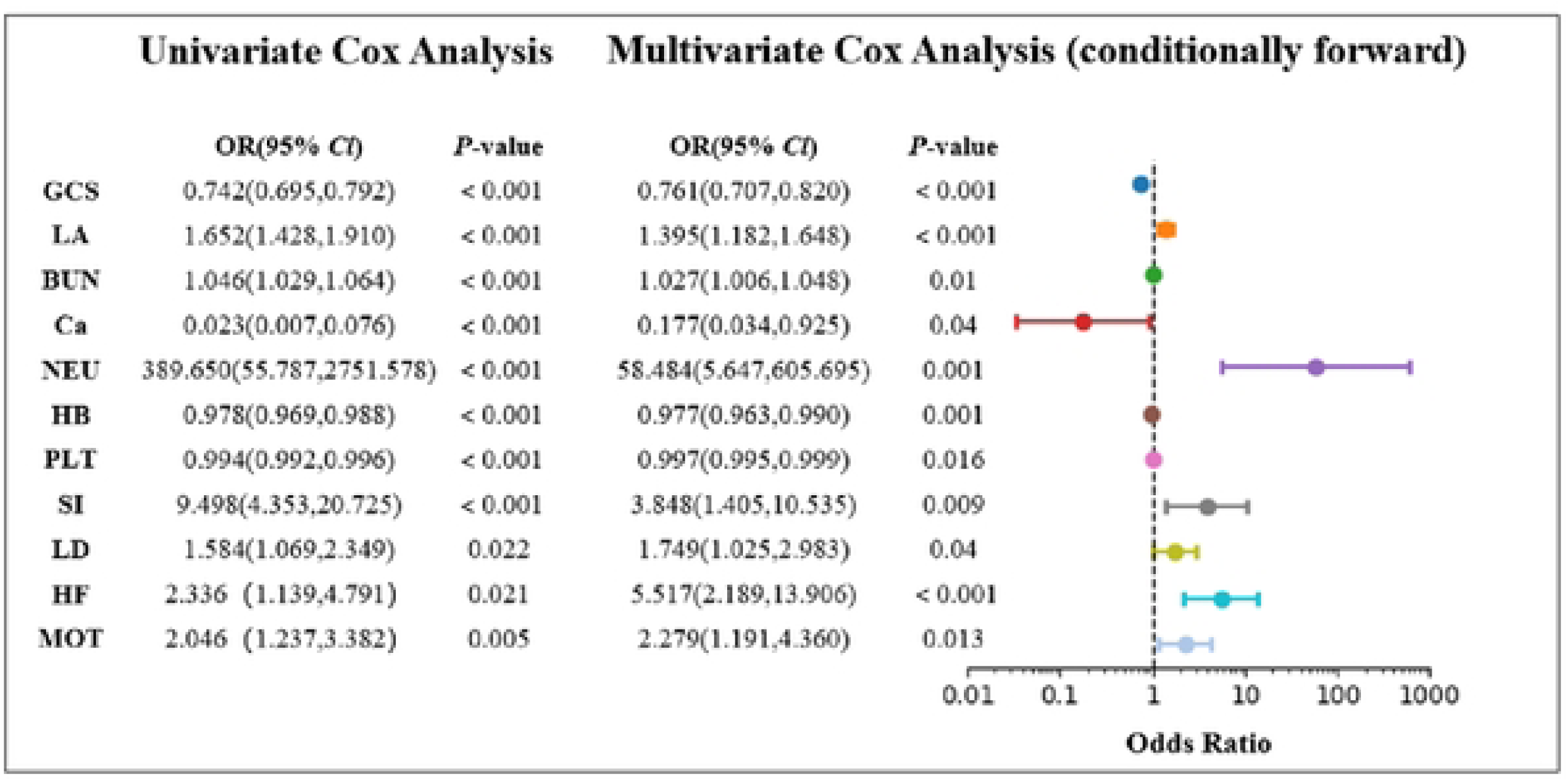
Screening of candidate rapid test features based on knowledge distillation. GCS, Glasgow Coma Scale; LA, Lactic acid; BUN, Blood urea nitrogen; Ca, Calcium; NEU, Neutrophil; HB, Hemoglobin; PLT, Platelet; MSI, Modified Shock Index; LD, Liver disease; HF, Heart failure; MOT, Metastasis of tumor.

On this basis, two senior emergency physicians added and removed features. First, hematological indicators were removed. Then, referring to PRS, AIMS-65, and GBS, indicators such as gender, age, heart rate, systolic blood pressure, and comorbidities were added. This formed 14 candidate features. Subsequently, heart rate was excluded via collinearity diagnosis (tolerance 0.158, VIF 6.348). Finally, combined with univariate and multivariate Logistic regression analysis results, comorbid ischemic heart disease and liver failure were excluded. Their Odds Ratios (OR) were both less than 1, appearing as protective factors. This is inconsistent with clinical experience. The reason may be that patients with chronic diseases discovered AUGIB risk factors and received interventions during regular treatment. Also, insufficient sample size causing data distribution bias cannot be excluded. Ultimately, 11 key features were confirmed as inputs for the rapid prediction model: Gender, Age, GCS, SBP, LD, Renal failure, HF, UGT, MOT, Shock, and SI (see supporting information S1 and S2). The preprocessed dataset was divided into a training cohort (480 cases) and a test cohort (121 cases) using stratified random sampling with an 8:2 ratio. The training cohort was oversampled using the Adaptive Synthetic Sampling Approach for Imbalanced Learning (ADASYN) [31] algorithm to achieve class balance, increasing the sample size to 716 cases.

### Evaluation metrics

Model performance was evaluated from three main aspects. (1) Discrimination: AUC served as the core evaluation metric, alongside Accuracy, Precision, Recall, and F1-score. (2) Calibration: The Brier Score (BS) was used for evaluation. Additionally, Calibration Curves were plotted for visual analysis. (3) Clinical utility and interpretability: Decision Curve Analysis was used to evaluate the Net Benefit at different threshold probabilities. SHAP values were introduced for interpretability analysis.

### Construction of teacher-student model cluster

To explore the performance of different algorithm architectures in distillation tasks, this study constructed a model cluster covering 7 mainstream ML algorithms. These included Extreme Gradient Boosting (XGBoost), Random Forest (RF), Gradient Boosting Machine (GBM), Logistic Regression (LR), Support Vector Machine (SVM), K-Nearest Neighbors (KNN), and Light Gradient Boosting Model (LightGBM).

### Statistical analysis

Data processing and statistical analysis were performed using SPSS 27.0. Normally distributed continuous variables were expressed as mean ± standard deviation (X^-^±s), and comparisons between groups were conducted using the t-test. Non-normally distributed continuous variables were expressed as median (interquartile range) (M (P25, P75)), and group comparisons were performed using the Mann-Whitney U test. Categorical variables were expressed as percentages (%), and comparisons were made using the Chi-square test or Fisher’s exact test. Statistical significance was defined as \**P* < 0.05.

### Ethics statement

This study was approved by the Ethics Committee of the First Medical Center of Chinese PLA General Hospital (Approval No. S2025-264-01), and informed consent was waived.

## Results

### Data baseline characteristics

A total of 601 patients were included in this study, with baseline characteristics detailed in Table 2. There were 430 cases in the favorable prognosis group and 171 cases in the adverse prognosis group. Overall, both groups were predominantly male (70.2% in the good prognosis group and 62.0% in the poor prognosis group). Among all candidate features, 36 features—including GCS, LA, and TBIL,—showed statistically significant differences between the two groups (\**P* < 0.05). The median values of 20 features, such as HR, LA, and AST, were higher in the poor prognosis group than in the good prognosis group. Conversely, the median values of 9 features, such as BE, ALB, and Ca, were significantly lower. Meanwhile, the PRS, GBS, AIMS-65, and MEWS scores were significantly higher in the poor prognosis group compared to the good prognosis group.

**Table 2.** Comparison of baseline characteristics in the favorable prognosis and adverse prognosis group.

| **Demographic characteristics** | **Favorable prognosis group,n=430** | **Adverse prognosis group,n=171** | **Z/χ²** | ***P* value** |
|---|---|---|---|---|
| **Demographic Information** |  |  |  |  |
| Gender |  |  | 3.814 | 0.051 |
| Male,n(%) | 302(70.2) | 106(62.0) |  |  |
| Female,n(%) | 128(29.8) | 65(38.0) |  |  |
| Age(yd) | 64.500(53.750,74.000) | 66.000(61.000,72.000) | -1.747 | 0.081 |
| **Vital Signs** |  |  |  |  |
| T(°C) | 36.500(36.400,36.700) | 36.500(36.400,36.800) | -1.535 | 0.125 |
| HR(BPM) | 78.000(68.000,92.000) | 92.000(75.000,107.00) | -5.418 | <0.001 |
| RR(BPM) | 19.000(17.000,20.000) | 19.000(17.000,20.000) | -0.206 | 0.837 |
| SBP(mmHg) | 118.000(103.000,133.000) | 113.000(100.000,130.000) | -1.780 | 0.075 |
| DBP(mmHg) | 66.000(60.000,75.000) | 65.000(59.000,72.000) | -1.820 | 0.069 |
| **Blood Test Results** |  |  |  |  |
| PH | 7.450(7.420,7.480) | 7.443(7.390,7.500) | -1.073 | 0.283 |
| PO2(mmHg) | 124.000(92.750,156.250) | 125.000(88.000,154.000) | -0.176 | 0.860 |
| PCO2(mmHg) | 33.000(29.000,37.000) | 30.000(25.000,37.000) | -2.935 | 0.003 |
| BE(mmol/L) | -0.300(-2.700,2.400) | -2.100(-6.000,1.100) | -4.106 | <0.001 |
| LA(mmol/L) | 1.000(0.700,1.500) | 1.700(1.100,3.100) | -8.956 | <0.001 |
| ALT(U/L) | 13.400(8.800,22.325) | 16.500(8.200,29.400) | -1.488 | 0.137 |
| AST(U/L) | 19.000(12.900,30.700) | 27.600(14.200,58.500) | -4.104 | <0.001 |
| ALB(g/L) | 31.100(27.675,34.300) | 26.600(24.400,30.100) | -8.343 | <0.001 |
| TBIL(umol/L) | 10.700(6.550,20.000) | 13.400(8.3000,27.200) | -2.896 | 0.004 |
| GLU(mmol/L) | 7.760(6.148,9.953) | 9.280(6.700,11.620) | -4.455 | <0.001 |
| BUN(mmol/L) | 6.885(4.298,10.790) | 11.810(7.370,17.600) | -6.835 | <0.001 |
| SCR(umol/L) | 74.950(59.425,96.850) | 91.600(58.400,169.100) | -3.371 | <0.001 |
| Ca(mmol/L) | 2.020(1.9300,2.120) | 1.960(1.850,2.050) | -6.070 | <0.001 |
| K(mmol/L) | 3.905(3.628,4.280) | 3.970(3.660,4.370) | -1.238 | 0.216 |
| Na(mmol/L) | 137.400(134.600,139.600) | 137.000(133.600,140.800) | -0.228 | 0.820 |
| AMY(U/L) | 51.300(35.300,81.350) | 52.400(29.000,89.800) | -0.317 | 0.751 |
| LIP(U/L) | 35.300(22.700,62.900) | 38.200(19.300,63.051) | -0.404 | 0.686 |
| NEU | 0.796(0.694,0.865) | 0.860(0.781,0.917) | -6.645 | <0.001 |
| WBC(10<sup>9</sup>/L) | 7.965(5.105,10.365) | 8.070(5.620,12.080) | -1.826 | 0.068 |
| HB(g/L) | 78.000(69.000,88.000) | 74.000(61.000,83.000) | -3.708 | <0.001 |
| PLT(10<sup>9</sup>/L) | 161.000(100.750,230.250) | 102.000(55.000,177.000) | -5.988 | <0.001 |
| HCT(%) | 0.228(0.199,0.264) | 0.214(0.180,0.246) | -3.544 | <0.001 |
| CRP(mg/dL) | 1.504(0.367,3.677) | 3.677(0.914,5.819) | -4.754 | <0.001 |
| IL－6(pg/mL) | 39.410(8.093,251.326) | 133.900(25.050,251.326) | -5.123 | <0.001 |
| PCT(ng/mL) | 0.367(0.113,3.274) | 2.210(0.378,3.274) | -6.525 | <0.001 |
| APTT(s) | 34.550(28.575,39.000) | 39.200(33.700,44.900) | -6.393 | <0.001 |
| PT(s) | 15.000(13.500,16.225) | 16.300(14.500,18.900) | -6.041 | <0.001 |
| INR | 1.245(1.110,1.693) | 1.440(1.210,2.150) | -4.107 | <0.001 |
| FIB(g/L) | 2.730(2.000,3.605) | 2.230(1.630,3.280) | -4.037 | <0.001 |
| D－Dimer(ug/mL) | 1.585(0.630,3.060) | 2.550(1.690,5.880) | -6.005 | <0.001 |
| L/A(mmol/g) | 0.031(0.021,0.049) | 0.064(0.039,0.131) | -9.757 | <0.001 |
| U/C | 0.083(0.600,0.119) | 0.109(0.076,0.166) | -4.901 | < 0.001 |
| **Primary Symptoms** |  |  |  |  |
| Hematemesis,n(%) | 174(40.5) | 101(40.9) | 0.011 | 0.916 |
| Syncope,n(%) | 53(12.3) | 22(12.9) | 0.033 | 0.857 |
| **Comorbidities and Underlying Diseases** |  |  |  |  |
| LD,n(%) | 97(22.6) | 54(31.6) | 5.292 | 0.021 |
| LF,n(%) | 70(16.3) | 22(12.9) | 1.100 | 0.294 |
| Renal failure,n(%) | 26(6.0) | 22(12.9) | 7.741 | 0.005 |
| HF,n(%) | 17(4.0) | 15(8.8) | 5.635 | 0.018 |
| Respiratory failure,n(%) | 6(1.4) | 12(7.0) | 13.311 | < 0.001 |
| IHD,n(%) | 18(4.2) | 5(2.9) | 0.529 | 0.467 |
| DM,n(%) | 95(22.1) | 32(18.7) | 0.838 | 0.360 |
| HTN,n(%) | 161(37.4) | 83(48.5) | 6.246 | 0.012 |
| CHD,n(%) | 71(16.5) | 22(12.9) | 1.244 | 0.265 |
| UGT,n(%) | 98(22.8) | 51(29.8) | 3.246 | 0.072 |
| OT,n(%) | 36(6.4) | 20(11.7) | 1.600 | 0.206 |
| MOT,n(%) | 42(9.8) | 31(18.1) | 8.015 | 0.005 |
| Shock,n(%) | 47(10.9) | 42(24.6) | 18.020 | < 0.001 |
| **Severity of Illness** |  |  |  |  |
| GCS | 15(15,15) | 13(7,15) | -12.659 | < 0.001 |
| SI | 0.655(0.566,0.804) | 0.779(0.611,0.945) | -5.162 | < 0.001 |
| Age－SI | 40.979(33.327,54.049) | 50.019(39.675,67.029) | -6.025 | < 0.001 |
| MSI | 0.921(0.793,1.111) | 1.074(0.873,1.360) | -5.234 | < 0.001 |
| PRS | 2(1,3) | 3(1,4) | -4.863 | < 0.001 |
| GBS | 9(7,11) | 12(10,13) | -8.532 | < 0.001 |
| AIMS－65 | 1(0,2) | 2(2,3) | -11.121 | < 0.001 |
| MEWS | 1(1,2) | 3(2,4) | -8.838 | < 0.001 |

Table notes: T, Temperature; HR, Heart rate; RR, Respiratory rate; SBP, Systolic blood pressure; DBP, Diastolic blood pressure; PO2, Arterial O2 pressure; PCO2, Arterial CO2 pressure; BE, Base excess; LA, Lactic acid; ALT, Alanine aminotransferase; AST, Aspartate aminotransferase; ALB, Albumin; TBIL, Total bilirubin; GLU, Glucose; BUN, Blood urea nitrogen; SCR, Serum creatinine; Ca, Calcium; K, Potassium; Na, Sodium; AMY, Amylase; LIP, Lipase; NEU, Neutrophil; WBC, White blood cell; HB, Hemoglobin; PLT, Platelet; HCT, Hematocrit; CRP, C-reactive protein; IL-6, Interleukin-6; PCT, Procalcitonin; APTT, Activated partial thromboplastin time; PT, Prothrombin time; INR, International normalized ratio; FIB, Fibrinogen; L/A, Lac/Albumin; U/C, Urea/Creatinine; LD, Liver disease; LF, Liver failure; HF, Heart failure; IHD, Ischemic heart disease; DM, Diabetes mellitus; HTN, Hypertension coronary heart disease; CHD, Coronary heart disease; UGT, Upper gastrointestinal tumor; OT, Other tumors; MOT, Metastasis of tumor.

### Model performance and visualization

In the Teacher-Student model cluster, AUC was used as the primary evaluation metric. Based on this metric, LightGBM was identified as the optimal model for both the teacher model and the rapid prediction model. Hyperparameter search was performed using Bayesian optimization. The optimal hyperparameter combination for the rapid prediction model is shown in supporting information S3. The LightGBM teacher model achieved an AUC of 0.867. Its accuracy, precision, recall, and F1 score were 0.818, 0.816, 0.702, and 0.755, respectively (Fig 3a). The LightGBM rapid prediction model achieved an AUC of 0.848. Its accuracy, precision, recall, and F1 score were 0.826, 0.750, 0.732, and 0.741, respectively (Fig 3b). In terms of probability prediction, the Brier scores were 0.148 and 0.128, respectively. Calibration curves (Fig 4) showed that both models were close to the ideal calibration line across probability intervals, indicating good accuracy and reliability. Decision curves (Fig 5) showed that the models had high net benefit over a wide range of probability thresholds, suggesting good clinical application value. Compared to the optimal teacher model, the AUC of the LightGBM rapid prediction model decreased by 0.014, maintaining good discrimination performance. The performance of each model is listed in Table 3.

**Fig 3.**
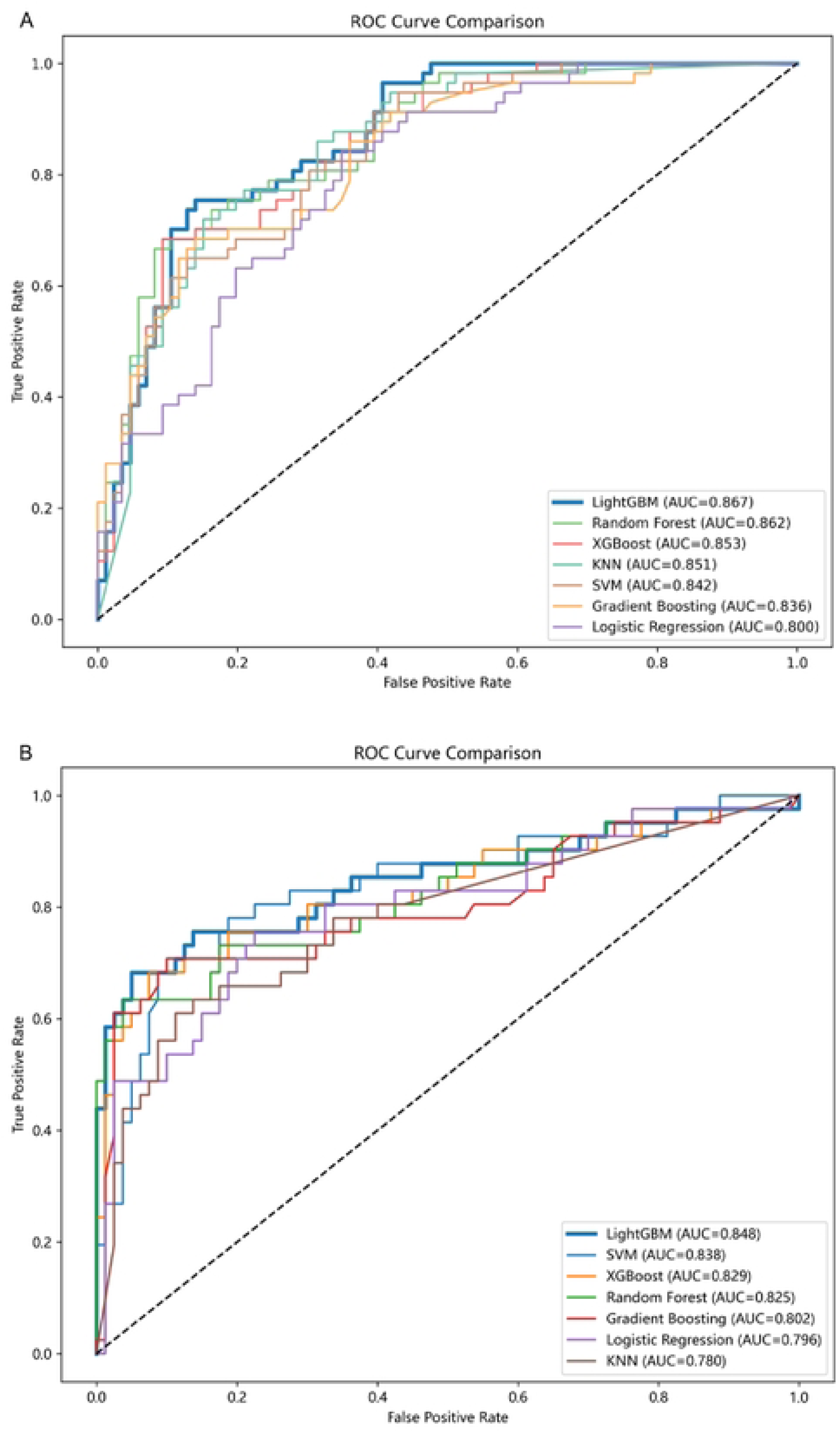
ROC curves. (A) Teacher model receiver operating characteristic curve. (B) The rapid prediction model receiver operating characteristic curve.

**Fig 4.**
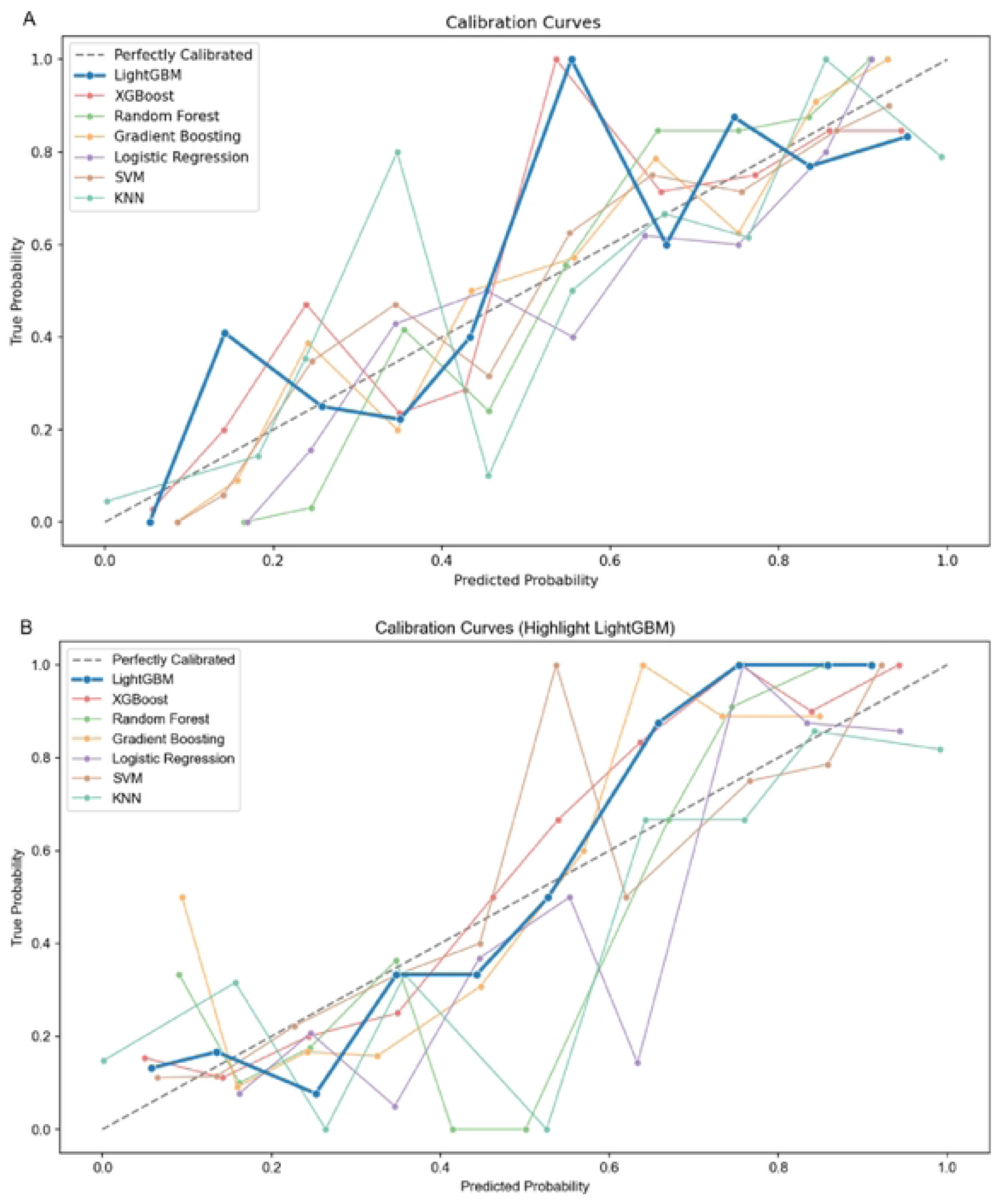
Calibration curves. (A) Teacher model calibration curve. (B) The rapid prediction model calibration curve.

**Fig 5.**
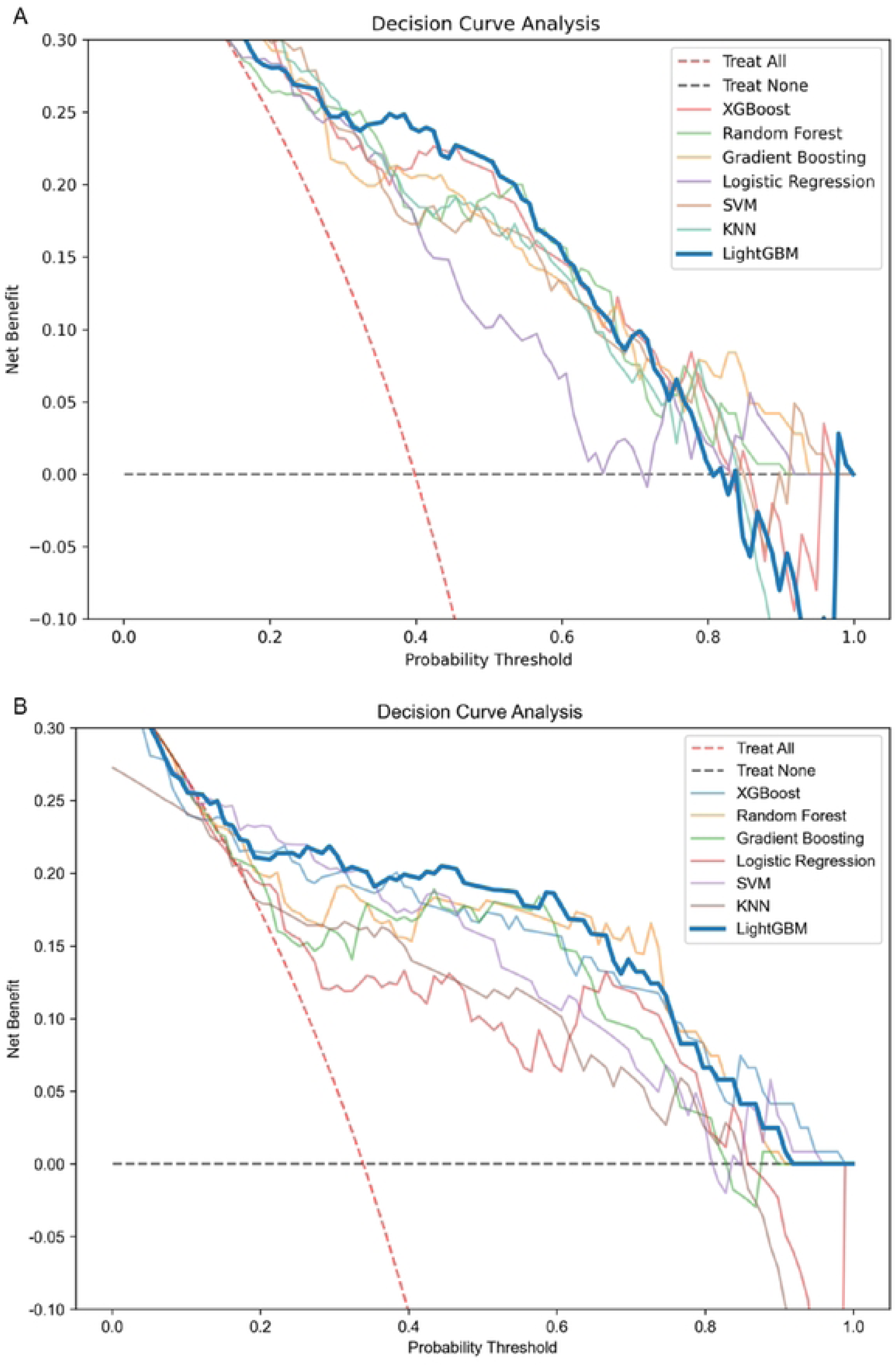
Decision curves. (A) Teacher model decision curve. (B) The rapid prediction model decision curve.

**Table 3.** Comparison of the main performance parameters of every models.

| **Model** | **AUC** | **Accuracy** | **Precision** | **Recall** | **F1-score** | **Brier** |
|---|---|---|---|---|---|---|
| XGBoost-T | 0.853 (0.792, 0.909) | 0.811 (0.748, 0.874) | 0.826 (0.714, 0.929) | 0.667 (0.545, 0.783) | 0.738 (0.637, 0.820) | 0.153 (0.117, 0.194) |
| RF-T | 0.862 (0.797, 0.917) | 0.790 (0.720, 0.853) | 0.755 (0.632, 0.865) | 0.702 (0.582, 0.820) | 0.727 (0.624, 0.811) | 0.162 (0.141, 0.184) |
| GBM-T | 0.836 (0.766, 0.897) | 0.776 (0.706, 0.839) | 0.778 (0.644, 0.889) | 0.614 (0.500, 0.742) | 0.686 (0.580, 0.778) | 0.160 (0.127, 0.193) |
| LR-T | 0.800 (0.730, 0.867) | 0.706 (0.636, 0.783) | 0.627 (0.508, 0.750) | 0.649 (0.519, 0.774) | 0.638 (0.532, 0.734) | 0.184 (0.156, 0.213) |
| SVM-T | 0.842 (0.772, 0.898) | 0.776 (0.706, 0.839) | 0.791 (0.666, 0.900) | 0.596 (0.471, 0.729) | 0.680 (0.568, 0.776) | 0.161 (0.129, 0.194) |
| KNN-T | 0.851 (0.784, 0.909) | 0.783 (0.713, 0.839) | 0.717 (0.600, 0.825) | 0.754 (0.642, 0.864) | 0.735 (0.641, 0.814) | 0.161 (0.122, 0.205) |
| **LightGBM-T** | **0.867 (0.803, 0.921)** | **0.818 (0.748, 0.881)** | **0.816 (0.700, 0.920)** | **0.702 (0.586, 0.820)** | **0.755 (0.660, 0.842)** | **0.148 (0.109, 0.190)** |
| XGBoost-S | 0.829 (0.733, 0.914) | 0.802 (0.727, 0.868) | 0.707 (0.555, 0.841) | 0.707 (0.562, 0.840) | 0.707 (0.575, 0.810) | 0.136 (0.093, 0.183) |
| RF-S | 0.825 (0.723, 0.906) | 0.760 (0.686, 0.835) | 0.625 (0.488, 0.761) | 0.732 (0.577, 0.861) | 0.674 (0.552, 0.778) | 0.139 (0.109, 0.174) |
| GBM-S | 0.802 (0.704, 0.894) | 0.686 (0.603, 0.769) | 0.525 (0.400, 0.644) | 0.780 (0.645, 0.900) | 0.627 (0.506, 0.726) | 0.150 (0.120, 0.186) |
| LR-S | 0.796 (0.697, 0.887) | 0.570 (0.487, 0.661) | 0.430 (0.317, 0.540) | 0.829 (0.714, 0.935) | 0.567 (0.453, 0.667) | 0.172 (0.142, 0.203) |
| SVM-S | 0.838 (0.753, 0.911) | 0.777 (0.702, 0.851) | 0.640 (0.500, 0.765) | 0.780 (0.636, 0.895) | 0.703 (0.583, 0.796) | 0.142 (0.100, 0.182) |
| KNN-S | 0.780 (0.687, 0.865) | 0.744 (0.669, 0.818) | 0.614 (0.477, 0.750) | 0.659 (0.500, 0.800) | 0.635 (0.507, 0.742) | 0.176 (0.121, 0.232) |
| **LightGBM-S** | **0.848 (0.757, 0.920)** | **0.826 (0.760, 0.893)** | **0.750 (0.605, 0.889)** | **0.732 (0.595, 0.857)** | **0.741 (0.620, 0.839)** | **0.128 (0.089, 0.169)** |

Table notes: T represents the Teacher Model; S represents the Student Model, which is the Rapid Prediction Model.

### Model, which is the Rapid Prediction Model

The SHAP algorithm was used to interpret the LightGBM rapid prediction model. The SHAP summary plot (Fig 6) shows the top three features contributing to the model prediction. They are GCS, Age, and Gender, in that order. GCS was negatively correlated with poor prognosis. This means a higher GCS score indicates a lower risk of poor prognosis. Age was positively correlated with poor prognosis. Older age indicated a higher risk. Female patients had a higher risk of poor prognosis than male patients. The pie chart in the lower right corner shows the contribution proportion of each data type. Comorbidities accounted for the highest proportion (33.8%), followed by simple scoring indicators (33.0%). Vital signs accounted for the smallest proportion (6.3%).

**Fig 6.**
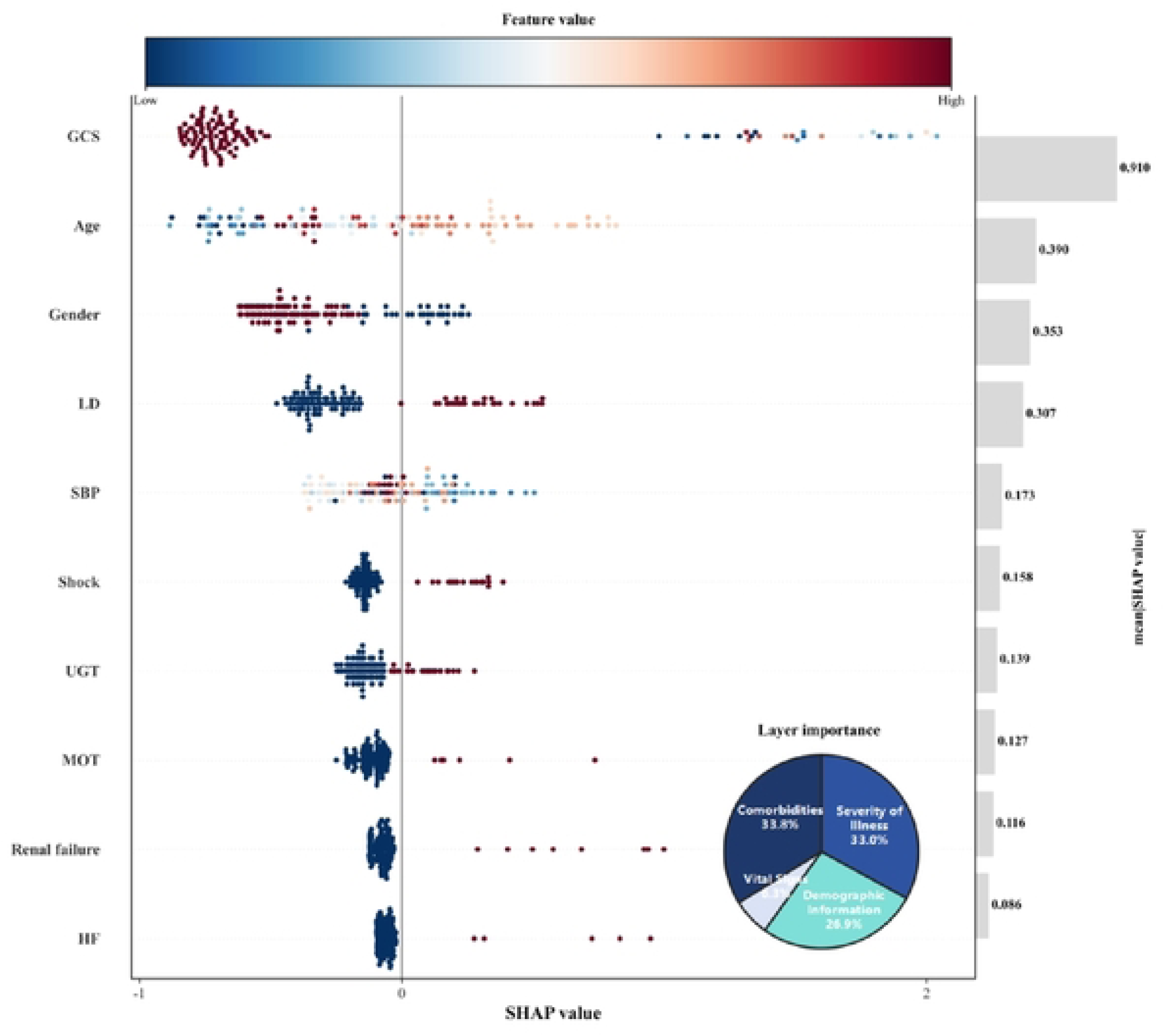
SHAP summary plot of the optimal rapid prediction model LightGBM. Features are arranged by importance from top to bottom. GCS made the greatest contribution, while HF made the least. Each dot represents a sample. Colors range from blue to red, indicating feature values from low to high. The distance of the dot from the zero line reflects the direction and magnitude of the feature’s impact. The left side of the zero line indicates a negative effect, while the right side indicates a positive effect.

### Comparison of clinical risk assessment systems and the optimal rapid prediction model

The performance of 7 clinical scoring systems and the LightGBM rapid prediction model was compared on the test cohort (detailed in Table 4). In the clinical scoring subgroup, using AUC as the main metric, GBS performed best (AUC 0.754). Accuracy was highest for the AIMS-65 score at 0.727. Precision was highest for the MEWS scoring system at 0.595. Recall was highest for the GBS scoring system at 0.781, indicating a lower missed diagnosis rate. The Delong test was used to compare AUC differences among models (detailed in Table 4). Results showed that LightGBM’s predictive ability was significantly better than PRS, SI, Age-SI, and MSI (\**P* < 0.05). Compared to GBS, AIMS-65, and MEWS, although the difference was not statistically significant, LightGBM performed better in accuracy and precision, showing good clinical application potential. In terms of timeliness, LightGBM relies only on immediate indicators such as vital signs and comorbidities. This is consistent with the design concepts of PRS, SI, Age-SI, MSI, and MEWS, making it suitable for rapid emergency assessment. In contrast, GBS and AIMS-65 rely on hematological indicators such as BUN, HB, ALB, and INR, which require longer assessment time.

**Table 4.** Performance comparison of clinical risk assessment systems and the rapid prediction model LightGBM.

| **Scoring Systems** | **AUC（95%CI）** | **Bonferroni-corrected *P*-value** | **Accuracy** | **Precision** | **Recall** | **F1-score** | **Optimal threshold** | **Included Hematological Indicators** |
|---|---|---|---|---|---|---|---|---|
| the Rapid Prediction Model LightGBM | **0.848 (0.757, 0.920)** | - | **0.826** | **0.750** | **0.732** | **0.741** | 0.45 | - |
| PRS | 0.592(0.504, 0.680) | 0.024 | 0.678 | 0.546 | 0.293 | 0.381 | 4 | - |
| **GBS** | **0.754(0.676, 0.831)** | **1.000** | **0.686** | **0.525** | **0.781** | **0.628** | **10** | **BUN、HB** |
| AIMS－65 | 0.752(0.674, 0.830) | 1.000 | 0.727 | 0.577 | 0.732 | 0.645 | 2 | ALB、INR |
| MEWS | 0.717(0.636, 0.798) | 0.643 | 0.719 | 0.595 | 0.537 | 0.564 | 3 | - |
| SI | 0.549(0.460, 0.639) | 0.006 | 0.587 | 0.421 | 0.585 | 0.490 | 0.70 | - |
| Age－SI | 0.576(0.487, 0.664) | 0.015 | 0.620 | 0.449 | 0.537 | 0.489 | 44.79 | - |
| MSI | 0.580(0.492, 0.669) | 0.017 | 0.612 | 0.438 | 0.512 | 0.472 | 1.05 | - |

## Discussion

Based on the emergency setting, this study focused on the acute onset and rapid progression of AUGIB. To meet the dual need for rapid and accurate assessment of critically ill patients, we prioritized timeliness and constructed a rapid prediction model for 7-day adverse prognosis in AUGIB patients using a teacher-student model. Results showed that the rapid prediction model (student model) performed best with LightGBM. The AUC was 0.848, accuracy was 0.826, precision was 0.750, recall was 0.732, F1 score was 0.741, and Brier score was 0.128. This model relies on immediate indicators such as vital signs and comorbidities, so the assessment takes a short time. In terms of discrimination, the LightGBM rapid prediction model was significantly better than PRS, SI, Age-SI, and MSI. Compared with GBS, AIMS-65, and MEWS, although there was no statistical difference, it showed clear advantages in accuracy and precision. Meanwhile, compared with the optimal teacher model covering all features, its AUC decreased by only 0.014, maintaining good discrimination performance.

The first 7 days after AUGIB onset is the active phase. During this stage, bleeding is active, rebleeding risk is high, and disease progresses rapidly. Serious complications such as shock and multiple organ failure are likely. This is a critical window for clinical intervention. This study set the observation window at 7 days [32] to address key challenges in emergency AUGIB treatment. Therefore, poor prognosis was defined as rebleeding, all-cause death, and serious events such as emergency surgical or interventional treatment after medical and endoscopic therapy failure. The goal was to assist emergency decision-making and allocate limited resources, including emergency staff attention, ICU bed preparation, and resuscitation supplies. We also considered that doctor decisions might be influenced by external factors. For instance, studies suggest endoscopy should be completed within 24 hours or even less time [33,34], and clinical transfusion requires considering blood bank resources. Therefore, objective outcomes such as resuscitation and ICU transfer were included as observation endpoints to enhance reliability.

SHAP analysis showed that GCS, Age, and Gender had the greatest contribution. GCS is a recognized indicator for assessing consciousness. Lower scores reflect poorer consciousness and more severe illness. Its high contribution weight aligns with clinical experience, as doctors often make initial judgments based on consciousness. In addition, the value of GCS in predicting poor prognosis in patients with trauma [35,36] and acute pancreatitis [37] has been verified. Elderly patients have been confirmed to be closely related to poor prognosis due to frailty [38] and decreased immunity [39]. This is consistent with our findings. Regarding gender, previous literature indicates that ANVUGIB patients are mostly males aged 60 to 65 [40]. However, our SHAP analysis showed that female patients were more prone to poor prognosis. This may be related to the interaction between gender and other features. Other interpretable clinical parameters, such as comorbid liver disease, were introduced into the model and played an important role. This lays a foundation for the promotion of the model.

The advantage of this study lies in the introduction of a teacher-student knowledge distillation strategy. This strategy effectively achieved implicit knowledge transfer across feature spaces. The soft labels from the teacher model carry richer information than discrete hard labels. This is known as “dark knowledge.” They also serve as a strong regularization method. They guide the student model to reconstruct complex relationships among indicators in a limited feature space. This helps the student model learn prediction logic without relying on laboratory data. The constructed LightGBM rapid prediction model significantly improved timeliness while maintaining good performance. It effectively balanced the conflict between time sensitivity and information integrity in emergency assessment. This provides decision support for rapid patient stratification in high-throughput emergency settings. This design aligns with the concept of triage in mass casualty incidents [41]. It emphasizes identifying the most urgent patients quickly under limited resources. This helps allocate resources scientifically and intervene early. Previous studies show that simple scoring systems based on easily accessible parameters have good predictive value for AUGIB prognosis, such as MEWS ^42^, SI, MSI, and Age-SI ^43^. Our model inherits this core idea of rapid assessment using immediately available parameters. It also improves performance through knowledge distillation. In addition, the rapid assessment feature supports dynamic monitoring. This is valuable for AUGIB patients with rapid disease progression.

This study has the following limitations. The single-center design and limited sample size limited the external generalization ability of the model. It needs further verification with multi-center data. The retrospective design led to a lack of quantitative assessment of the model’s timeliness. Past medical history and medication use were not included in the analysis due to incomplete data acquisition. This may have an impact on model performance.

The rapid prediction model based on knowledge distillation constructed in this study showed better predictive performance for 7-day adverse prognosis in AUGIB patients than existing clinical scoring systems. It demonstrated good application potential in the emergency setting of this center. It is also expected to be applied to resource-limited scenarios such as pre-hospital care and community health centers.

## Data Availability

The datasets generated and/or analysed during the current study are not publicly available due to privacy and ethical restrictions but are available from the corresponding author on reasonable request and after approval by the institutional review board of the requester's institution.

## Acknowledgements

The authors have no acknowledgements to declare.

## Supporting information

**S1 Table.**
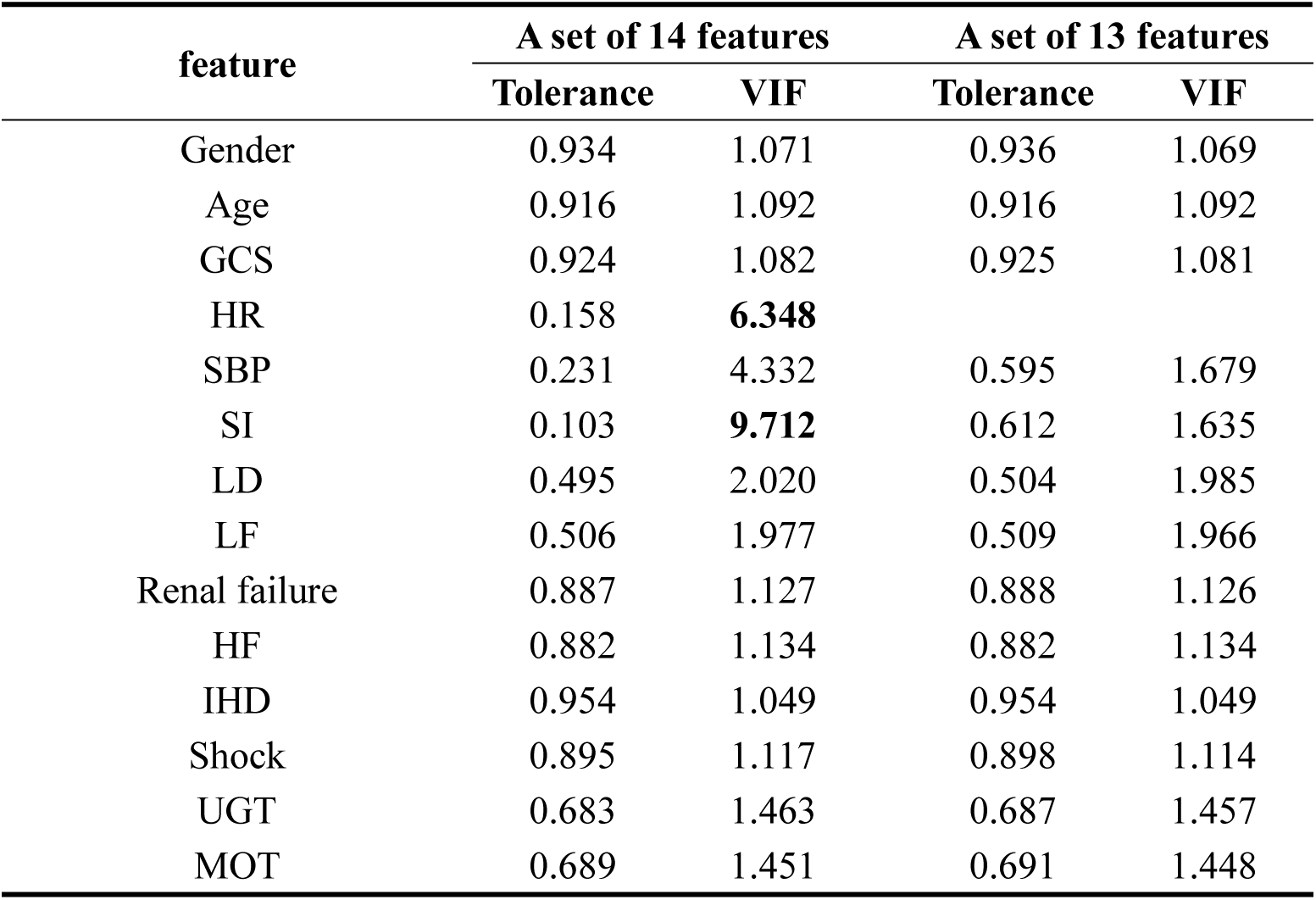
Collinearity analysis results of candidate features for rapid prediction.

**S2 Table.**
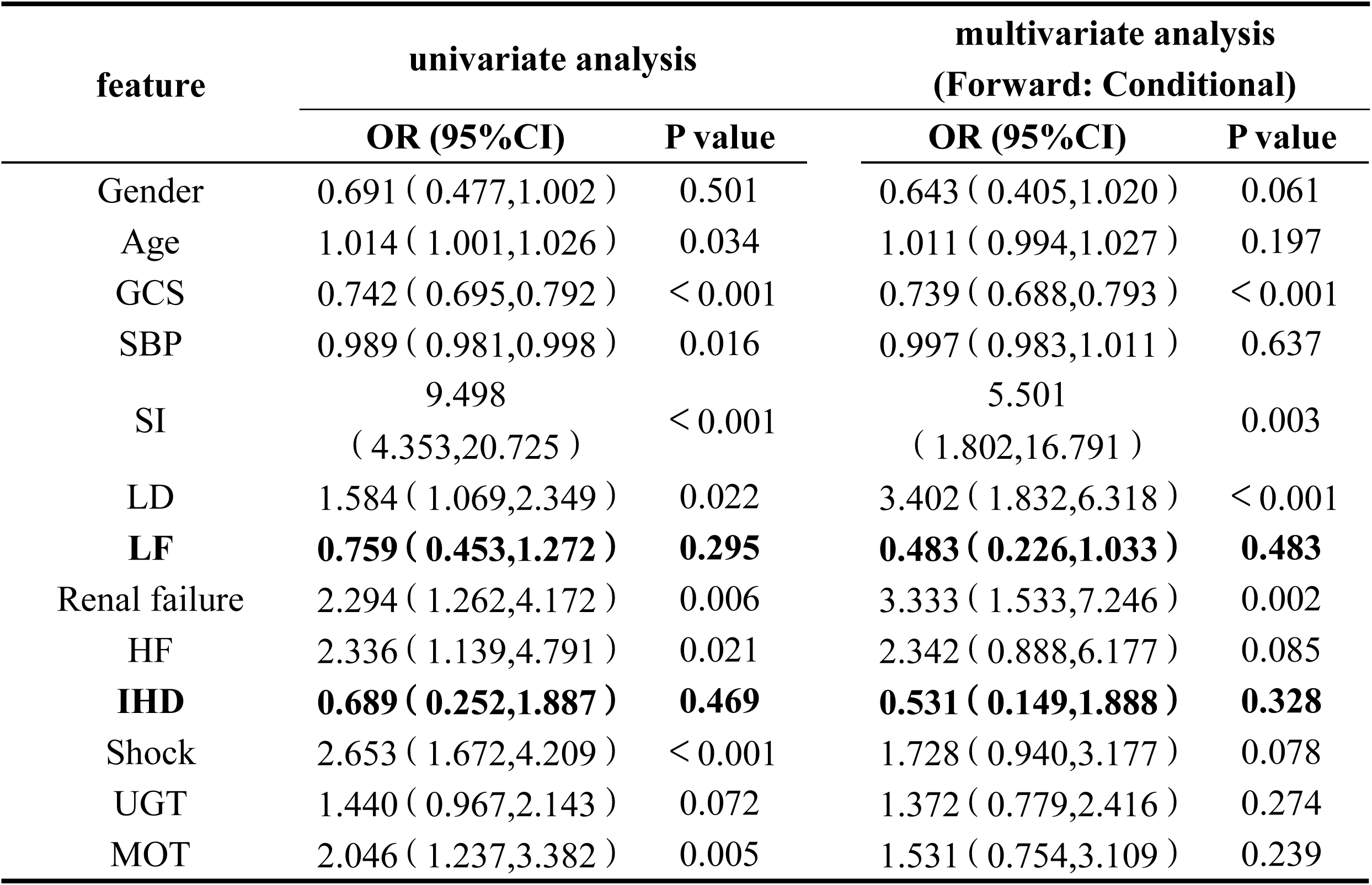
Results of univariate and multivariate analyses.

| feature | univariate analysis |  | multivariate analysis<br>(Forward: Conditional) |  |
|---|---|---|---|---|
|  | OR (95%CI) | P value | OR (95%CI) | P value |
| Gender | 0.691（0.477,1.002） | 0.501 | 0.643（0.405,1.020） | 0.061 |
| Age | 1.014（1.001,1.026） | 0.034 | 1.011（0.994,1.027） | 0.197 |
| GCS | 0.742（0.695,0.792） | ＜0.001 | 0.739（0.688,0.793） | ＜0.001 |
| SBP | 0.989（0.981,0.998） | 0.016 | 0.997（0.983,1.011） | 0.637 |
| SI | 9.498（4.353,20.725） | ＜0.001 | 5.501（1.802,16.791） | 0.003 |
| LD | 1.584（1.069,2.349） | 0.022 | 3.402（1.832,6.318） | ＜0.001 |
| **LF** | **0.759（0.453,1.272）** | **0.295** | **0.483（0.226,1.033）** | **0.483** |
| Renal failure | 2.294（1.262,4.172） | 0.006 | 3.333（1.533,7.246） | 0.002 |
| HF | 2.336（1.139,4.791） | 0.021 | 2.342（0.888,6.177） | 0.085 |
| **IHD** | **0.689（0.252,1.887）** | **0.469** | **0.531（0.149,1.888）** | **0.328** |
| Shock | 2.653（1.672,4.209） | ＜0.001 | 1.728（0.940,3.177） | 0.078 |
| UGT | 1.440（0.967,2.143） | 0.072 | 1.372（0.779,2.416） | 0.274 |
| MOT | 2.046（1.237,3.382） | 0.005 | 1.531（0.754,3.109） | 0.239 |

**S3 Table.** Optimal hyperparameters of the LightGBM rapid prediction model.

| **Hyperparameter** | **Value** |
|---|---|
| n_estimators | 200 |
| max_depth | 6 |
| num_leaves | 51 |
| learning_rate | 0.04 |
| subsample | 0.9241328468046672 |
| colsample_bytree | 0.85 |
| reg_lambda | 8.0 |
| reg_alpha | 1.5 |
| min_split_gain | 0.03 |
| min_child_samples | 25 |
| random_state | 42 |
| n_jobs | 1 |
| verbosity | -1 |

